# Comparative cardiovascular safety of romosozumab versus bisphosphonates in Japanese patients with osteoporosis A new-user, active comparator design with instrumental variable analyses

**DOI:** 10.1101/2024.03.14.24304284

**Authors:** Ryoji Tominaga, Tatsuyoshi Ikenoue, Ryosuke Ishii, Kakuya Niihata, Tetsuro Aita, Tadahisa Okuda, Sayaka Shimizu, Masataka Taguri, Noriaki Kurita

## Abstract

**Importance:** Romosozumab, a novel anti-osteoporotic agent, confers marked improvement in bone mineral density; however, its cardiovascular safety remains a concern.

**Objective:** To compare the cardiovascular safety of romosozumab to bisphosphonates in patients with osteoporosis.

**Design:** A cohort study using a new-user, active comparator design.

**Setting:** Medical facilities, including clinics and hospitals, visited by a wide range of populations in Japan that are covered by a commercial administrative claims database, collected from March 4, 2019 to August 31, 2021.

**Participants:** Japanese adults aged ≥40 years who were diagnosed with osteoporosis or experienced a fragility fracture. Those who received romosozumab or bisphosphonates after the commercialization of romosozumab started in Japan (March 4, 2019) were included.

**Exposure:** A new prescription of romosozumab or bisphosphonate (based on verification of a 180-day washout period).

**Main Outcomes and Measures:** The primary outcome was the incidence rate of cardiovascular disease (consisting of myocardial infarction and stroke) within one year of prescription. Cardiovascular disease was identified by algorithms with a combination of diagnosis, medical procedure, and drug codes. Facility-level prescription preference for romosozumab was used as an instrumental variable, defined as the proportion of romosozumab prescribed at the patient’s facility within 90 days prior to the index date.

**Results:** Of the 61,558 included prescriptions, 8,806 were for romosozumab and 52,752 were for bisphosphonates. The mean age of the romosozumab group was higher than that of the bisphosphonates group (80.5 vs. 78.3 years, respectively). The majority of patients were female (80.2 vs. 85.3%, respectively). The incidence of cardiovascular disease was 7.98 per 100 person-years for romosozumab versus 7.15 for bisphosphonates (unadjusted incidence rate ratio: 1.12 (95% confidence interval: 1.03-1.21)). An instrumental variable analysis using the two-stage residual inclusion method yielded a hazard ratio of 1.09 (95% confidence interval: 0.79-1.76) for romosozumab compared to bisphosphonates over one year.

**Conclusions and Relevance:** In this large observational study, there was no definitive evidence of increased risk of cardiovascular disease associated with romosozumab use compared with bisphosphonates in patients with osteoporosis. These findings alleviate clinicians’ excessive concerns about the potential cardiovascular safety of romosozumab in treatment decision-making for osteoporosis.

## Introduction

Osteoporosis is characterized by decreased bone density and compromised bone structure, and is particularly prevalent in the elderly.^1^ Treatments aimed at inhibiting bone resorption or promoting bone mineralization have been developed,^2–4^ as osteoporosis increases the risk of fragility fractures and negatively affects patients’ quality of life and overall survival.^5,6^

Romosozumab is a human monoclonal antibody with bone-forming and bone resorption-inhibiting effects. The efficacy of romosozumab was highlighted by the decreased vertebral fracture incidence and increased bone mineral density in an international phase III trial.^7^ However, increased incidence of cardiovascular disease (CVD) was linked to romosozumab in two clinical trials.^7,8^ Consequently, global safety alerts were issued, particularly for patients with CVD history; however, there is insufficient real-world evidence showing that romosozumab is associated with an increased CVD incidence compared to other osteoporosis drugs.

Pharmacovigilance studies using adverse event reporting databases in the United States and Japan indicated an increase in CVD with romosozumab compared to other medications.^9,10^ However, these findings were derived from adverse event cases and may suffer from bias related to unmeasured underlying disease and reporting bias.^11^ In addition, post-marketing surveillance of romosozumab by a pharmaceutical company lacks evidence from active comparator studies.^12^ Therefore, well-designed comparative studies involving new users of romosozumab and other osteoporosis medications are needed. The purpose of this study was to analyze the association of romosozumab and bisphosphonates with the development of CVD events among patients with osteoporosis. A new-user design was employed to address selection bias, and an instrumental variable analysis was used to address confounding by indication.

## Methods

### Database

We used the DeSC database, a commercially available administrative claims database (DeSC Healthcare Inc., Tokyo, Japan; https://desc-hc.co.jp/company). The database’s credibility and its representativeness of the Japanese population was validated by a comprehensive study.^13^ The following data were collected: individual IDs, fundamental medical information, medical institution files, diagnosis code based on the International Statistical Classification of Diseases and Related Health Problems, 10th Revision (ICD-10), detailed medical procedure files, pharmaceutical files, and medical practice detail file, as well as files containing the dates of these actions. The study dataset included data collected between April 1, 2014 and August 31, 2021. No approval from an ethics committee was required as the data were anonymized.

### Study design and subjects

This was a cohort study using a new-user, active comparator approach.^14,15^ Patients who met all of the following four criteria were included: (1) age ≥ 40 years; (2) ICD-10 code "osteoporosis with/without pathological fractures" or experiencing fragility fractures^16^; (3) new prescription of either romosozumab (WHO-ATC code: M05BX06) or bisphosphonate; and (4) prescription date later than March 4, 2019 (<u>eTable 1)</u>. Patients whose diagnosis code included "suspect" were excluded. The framework for the design details is presented in Figure 1. We followed the Strengthening the Reporting of Observational Studies in Epidemiology guidelines.

**Figure 1.**
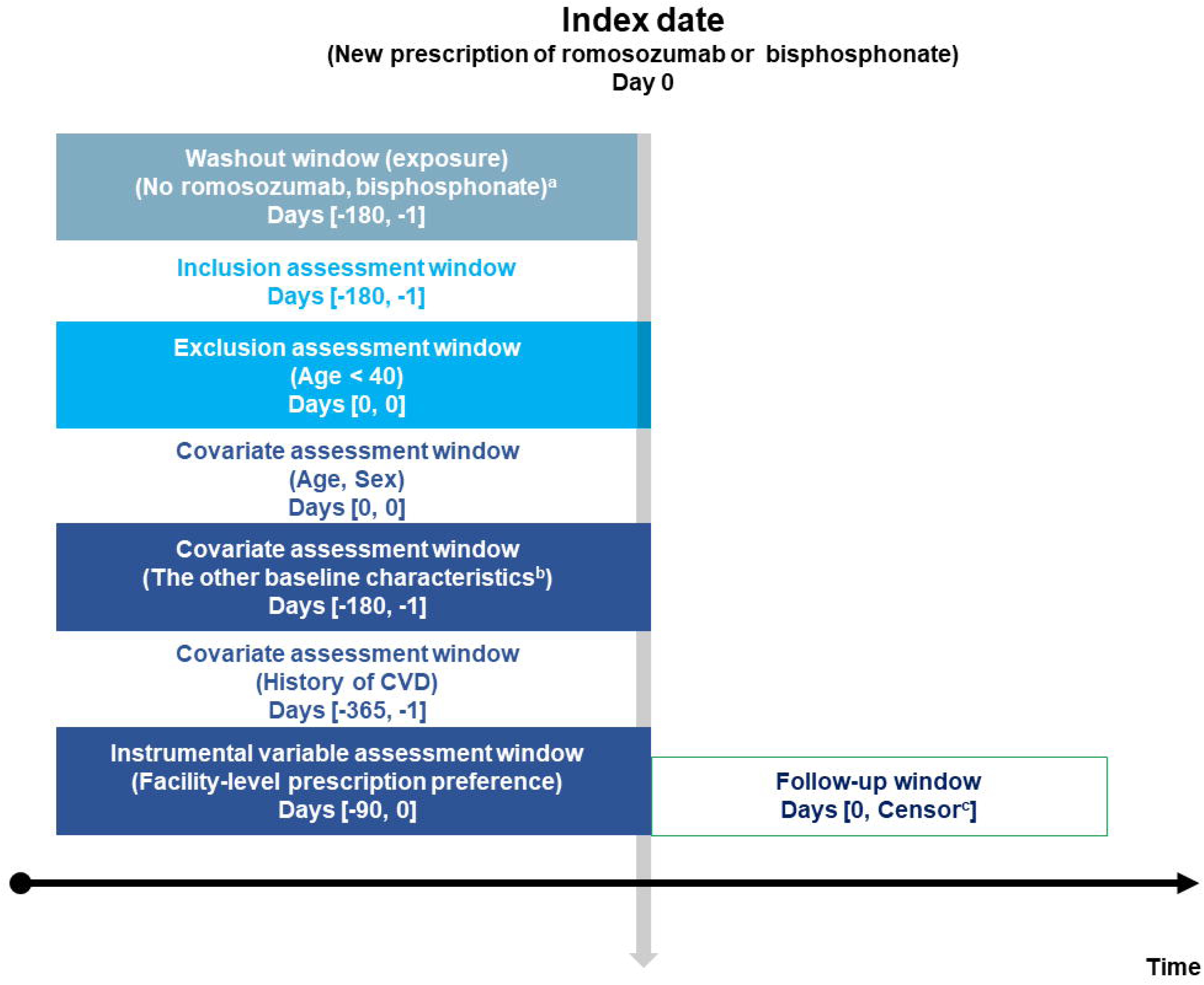
Framework for the design details of the present study. ^a^By the definition of 180-day washout, cases with <180 days of data due to a switch over or new enrollment of health insurance during this period were not included. ^b^Baseline characteristics included: diabetes, dyslipidemia, hypertension, chronic obstructive pulmonary disease, chronic kidney disease, rheumatoid arthritis, atrial fibrillation, the use of glucocorticoids. Censored at earliest of the defined outcomes, death, loss of the claim records (caused by loss of coverage under the three national health insurance programs, switch from health insurance to the public welfare system, or discontinuation of medical visit), non-persistence of the medications, end of the defined observation period (i.e., one or two-year), or end of the claim database (August 31, 2021) CVD = cardiovascular disease consisting of myocardial infarction or stroke (ischemic or hemorrhagic)

### Definition of exposure and comparator

The exposure and comparator were romosozumab and bisphosphonate, respectively. We chose bisphosphonate as the comparator since it is the most commonly administered osteoporosis drug; it was also chosen as the romosozumab comparator in two previous clinical trials.^7,8^ A list of the bisphosphonate codes indicated for osteoporosis in Japan is provided in <u>eTable 2</u>.

We defined "new use" as at least one filled prescription and no claims in a 180-day look-back period from the first dispensing date (i.e., index date).

The refill-gap approach was applied to determine the medication usage period.^14^ A medication was deemed persistent if the gap (i.e., the period between the last medication supply date and the date of the subsequent prescription) was ≤180 days, whereas a gap of >180 days was classified as non-persistent. The 180 days were based on the period of medication administration, accounting for drug clearance in the body, plus a grace period (twice the number of days of medication administration) accounting for delays in patient office visits. If the medication was switched to another anti-osteoporotic agent, that point was regarded as the end of the usage period.

### Definition of outcomes

The primary outcome was defined as a CVD event within 1 year of the index date. CVD event was defined as the first event of either myocardial infarction (MI) or stroke (cerebral infarction [CI] or cerebral hemorrhage [CH]). These definitions were determined after discussion and review with board-certified cardiologists and neurologists, with reference to previous studies that examined algorithms for valid identification of CVD events using Japanese claims databases.^17–20^ The algorithms were defined by the ICD-10 diagnosis code, plus a code indicating the specific treatment (medical procedure code or drug code [WHO-ATC code]), a code indicating hospitalization, and a code indicating imaging tests (for CI and CH only) (eMethods 1 and <u>eTable 3</u>).

The start of the at-risk period for the event was set to the aforementioned index date. The reason for specifying 1 year for the at-risk period is based on the fact that this is the duration of romosozumab administration according to the Japanese pharmaceutical product labeling. Censoring was defined as the earliest of the following events: end of 1-year follow-up, claims database end date (August 31, 2021), loss of claim records prior to the database end date (caused by loss of coverage under the three national health insurance programs, switch from health insurance to the public welfare system, or discontinuation of medical visit), non-persistence of the medications, or death.

In addition, a secondary outcome was defined as the incidence of a CVD event within 2 years from the index date. This was done because the prescription of romosozumab could be extended at the discretion of treating physicians. Furthermore, the secondary outcomes were defined as the incidence of MI, CI, and CH, respectively. At-risk periods for the three outcomes were defined as both within 1 and 2 years.

### Definition of covariates

A directed acyclic graph (DAG) was created to visualize the relationship among romosozumab or bisphosphonates use, cardiovascular events, and other relevant variables.^21^ The model was based on the best available evidence or, when evidence was not available, expertise in the field of bone metabolism. The final DAG model was reviewed and approved by four researchers (RT, TA, KN, NK; <u>e</u>Figure 1). Based on the DAG, the analysis established in the Dagitty web application (http://www.dagitty.net) were used to determine whether a given variable should be considered a confounder.^22^

Patient-level confounding factors including demographic characteristics (age and sex) and comorbidities were measured. These comorbidities included diabetes, dyslipidemia, hypertension, chronic obstructive pulmonary disease (COPD); chronic kidney disease (CKD); rheumatoid arthritis (RA); atrial fibrillation (AF); and history of MI, CI, or CH (<u>eTable 4)</u>.^23^ Glucocorticoid use, considering its association with bone metabolism, was considered if it was continuously used for more than 3 months (<u>eTable 5)</u>. These variables were identified during the look-back period, except for history of MI, CI, and CH, which was considered within 1 year, as this is the specified period in the pharmaceutical label; moreover, these were determined based on observations available up to 1 year prior to the index date.

Facility-level confounders included facility function and number of beds. Facility function and number of beds are postulated to reflect structures and processes of a facility’s medical care and to influence osteoporosis medication prescribing and incidence of patient CVD. These medical facility variables were extracted from the "medical institution file" data. The function of medical facilities was classified according to the Diagnosis Procedure Combination (DPC) medical institution group classification into non-DPC, university DPC, designated hospital DPC, and standard hospital. The DPC system is a Japanese reimbursement system used mainly at university hospitals and advanced medical care centers. ^24^ Billing at DPC-affiliated hospitals is determined based on the diagnosis and treatment provided to patients.

### Instrumental variable

The facility-level proportion of romosozumab prescriptions, which denominator was defined as the sum of romosozumab and bisphosphonates prescriptions within 90 days prior to the index date, was chosen as the instrumental variable.^25,26^ This variable was expected to be a strong predictor of the newly prescribed anti-osteoporotic. We assumed that facility-level prescription preferences did not correlate with CVD incidence other than through the pathway of choice of osteoporosis medications. However, the number of prescriptions included prescriptions to all users meeting eligibility criterion 2) (i.e., including not new prescriptions and those to patients <40 years old) as well as to new users meeting eligibility criteria 1) and 2). This was because we considered clinically sensible that physicians selected osteoporosis drugs for new patients by referring to the recent prescription history. The prescription proportion of romosozumab was calculated for each medical institution and for each month from the index date.

Then, prescription preference deciles were used as instrumental variables after excluding facility-level prescription proportions of zero and those with <5 prescriptions. The former exclusion criteria was to ensure the two-stage residual inclusion (2SRI) method was used by enabling the variation in unmeasured confounding to be explained by the residuals (eMethods 2).^27^ The latter exclusion criteria, as noted in a previous study, is the minimum number of overall facility cases to estimate the instrumental variable.^28^

### Statistical analysis

Data are presented using means and standard deviations for continuous variables and frequencies and percentages for categorical variables. The number of events and incidence rates for the primary and secondary outcomes were described, and the unadjusted incidence rate ratios (IRRs) of romosozumab versus bisphosphonates were estimated using Poisson regression models.

To examine the associations between new prescriptions of romosozumab and outcome rates, we employed an instrumental variable approach based on the 2SRI method.^27,29^ In the first stage, new romosozumab or bisphosphonate prescription was regressed according to the decile of prescription preference using a mixed-effects logistic model. All patient- and facility-level confounders were entered as independent variables. The residual was calculated as the difference between the model-predicted probability of prescribing romosozumab for individual patients and the actual romosozumab prescription rate. Nagelkerke’s pseudo-R2 was computed based on a logistic regression model as an approximation to assess the model fit.^30,31^

In the second stage, the outcomes and their observation periods were regressed on the prescription of romosozumab or bisphosphonate and the residuals using Cox proportional hazard models for multiple-recorded data. Standard errors were estimated by 2000-time bootstrapping with patients as cluster units.^27^ All the aforementioned confounders were entered as independent variables.

All analyses were performed independently by both a trained epidemiologist (NK) and a biostatistician (RI) using Stata version 17 (StataCorp, College Station, TX) or R (R Foundation for Statistical Computing, Vienna, Austria). Each result was cross-checked and verified by senior biostatisticians (TO and MT).

## Results

Of the 144,948 observations, 37,178 were prescribed at facilities with <5 prescriptions within 3 months, 46,104 were prescribed at facilities with a zero romosozumab prescription proportion, 93 were missing covariate data, and 15 were missing outcome data, leaving a total of 61,558 observations included in the final analysis. (Figure 2)

**Figure 2:**
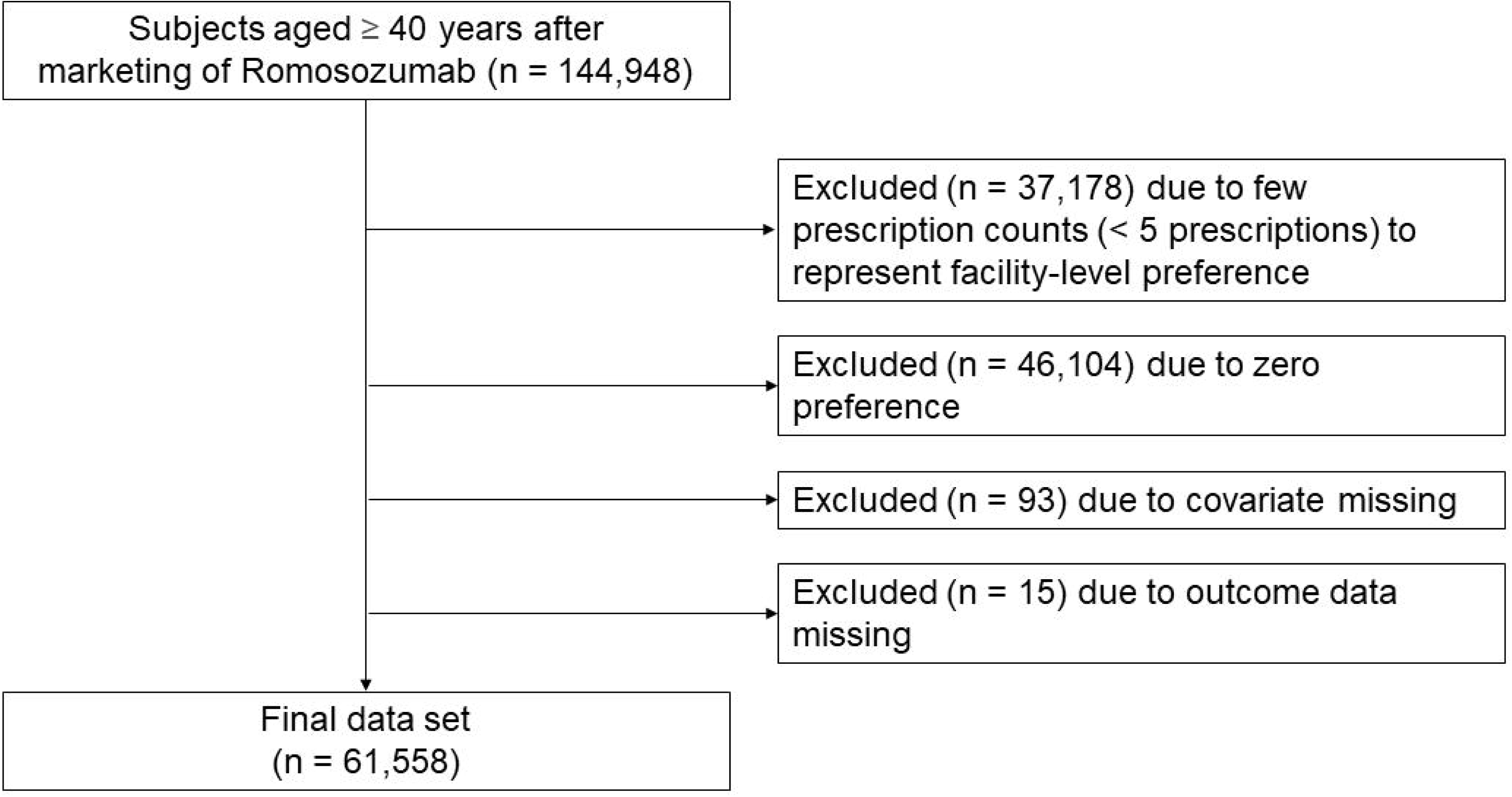
The flowchart of the subjects.

### Baseline characteristics of the participants

Baseline participant characteristics are shown in Table 1. Of the total observations, 6,1442 patients were unique and 116 had duplicate observations. There were 8,806 and 52,752 romosozumab and bisphosphonates observations from 8,774 and 52,552 unique patients, respectively.

**Table 1.**
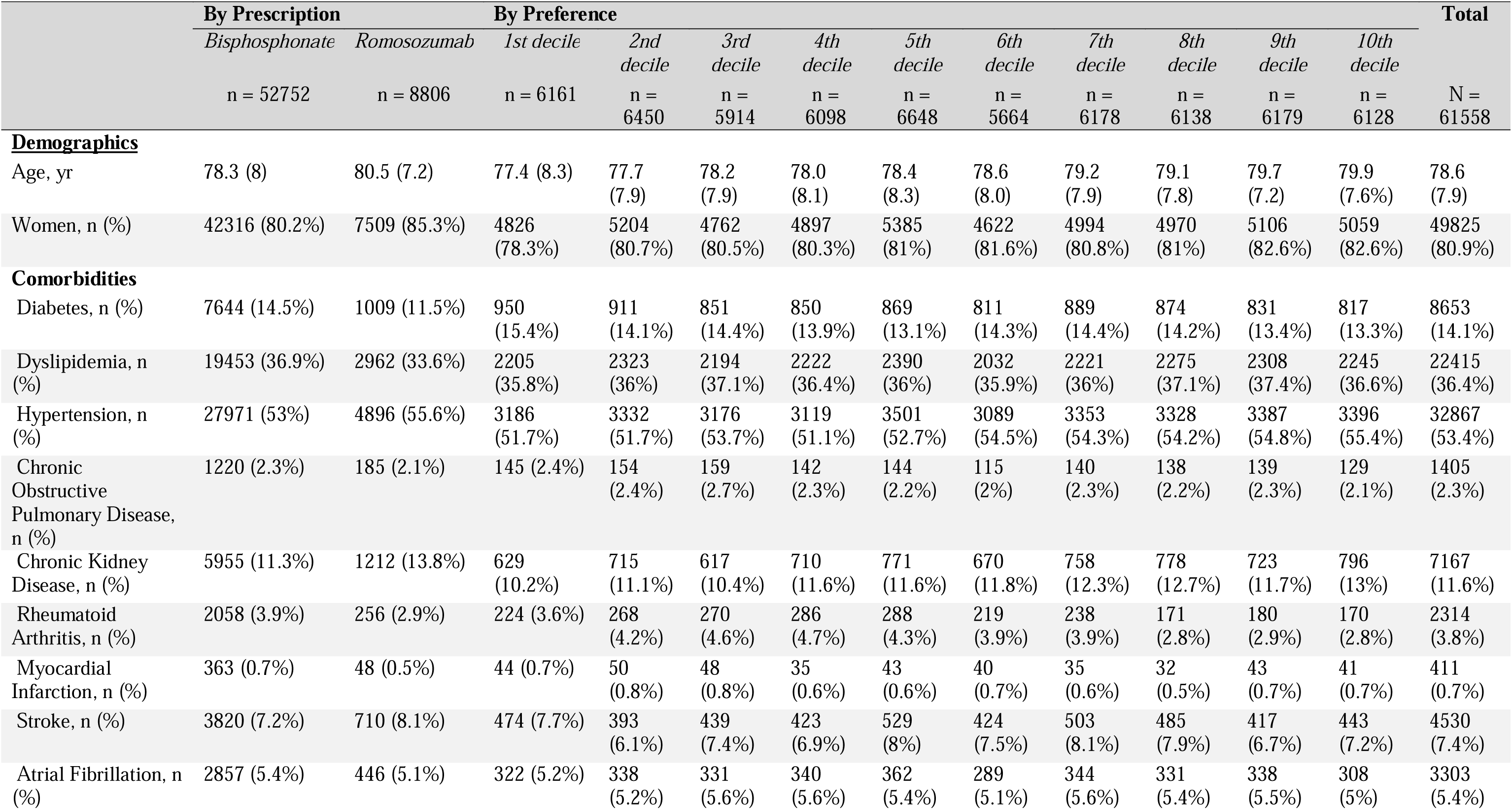
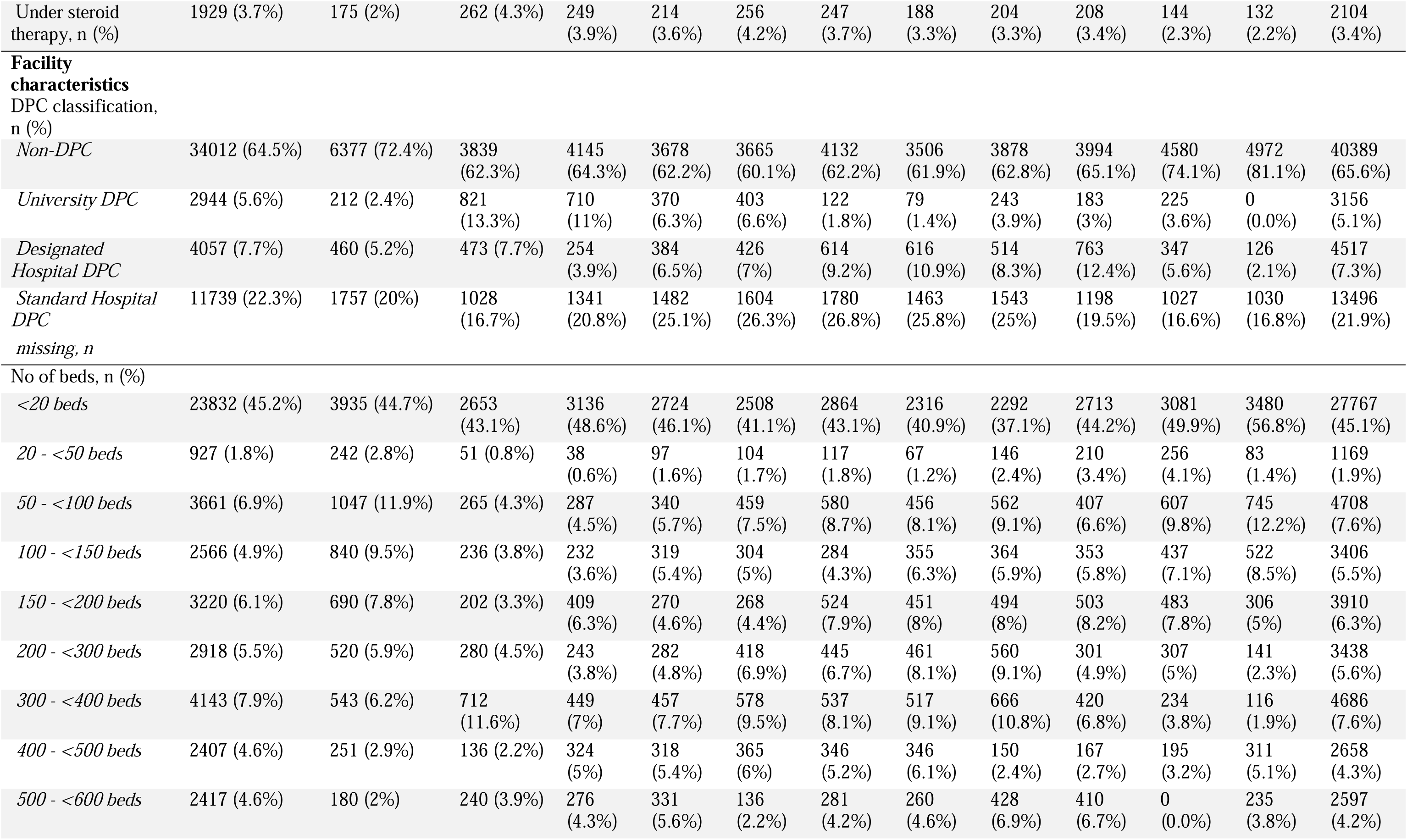
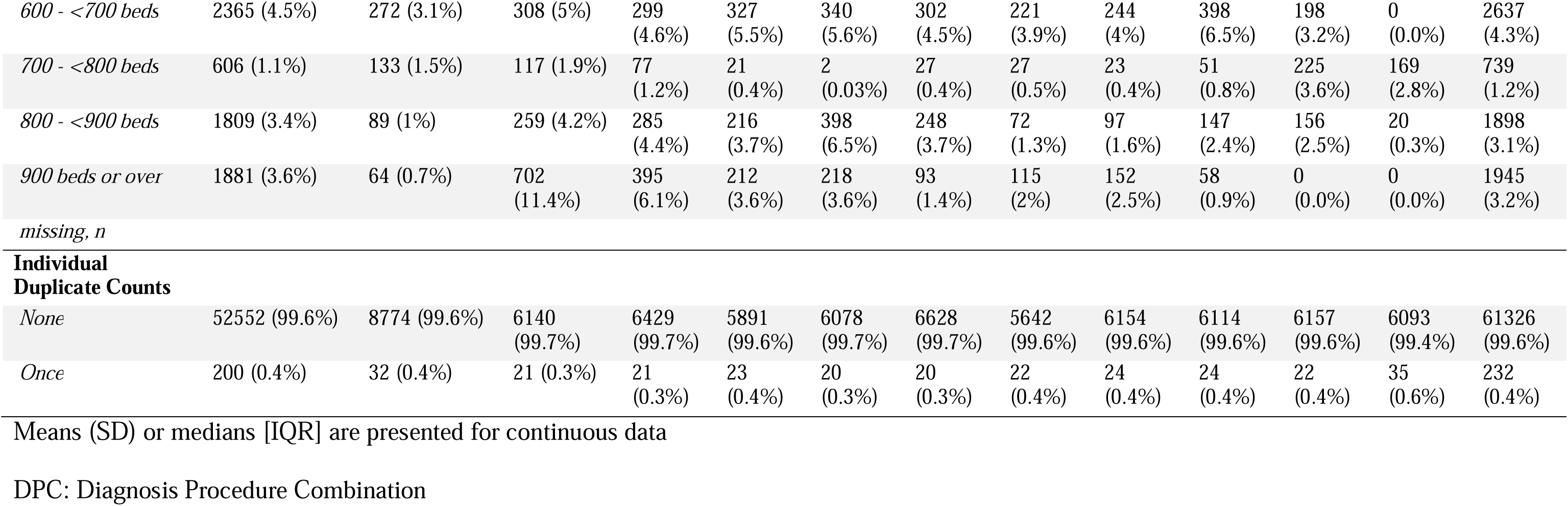
Characteristics of observations separated by prescription (N= 61,558)

|  | By Prescription |  | By Preference |  |  |  |  |  |  |  |  |  | Total |
| --- | --- | --- | --- | --- | --- | --- | --- | --- | --- | --- | --- | --- | --- |
|  | *Bisphosphonate* | *Romosozumab* | *1st decile* | *2nd decile* | *3rd decile* | *4th decile* | *5th decile* | *6th decile* | *7th decile* | *8th decile* | *9th decile* | *10th decile* |  |
|  | n = 52752 | n = 8806 | n = 6161 | n = 6450 | n = 5914 | n = 6098 | n = 6648 | n = 5664 | n = 6178 | n = 6138 | n = 6179 | n = 6128 | N = 61558 |
| **Demographics** |  |  |  |  |  |  |  |  |  |  |  |  |  |
| Age, yr | 78.3 (8) | 80.5 (7.2) | 77.4 (8.3) | 77.7 (7.9) | 78.2 (7.9) | 78.0 (8.1) | 78.4 (8.3) | 78.6 (8.0) | 79.2 (7.9) | 79.1 (7.8) | 79.7 (7.2) | 79.9 (7.6%) | 78.6 (7.9) |
| Women, n (%) | 42316 (80.2%) | 7509 (85.3%) | 4826 (78.3%) | 5204 (80.7%) | 4762 (80.5%) | 4897 (80.3%) | 5385 (81%) | 4622 (81.6%) | 4994 (80.8%) | 4970 (81%) | 5106 (82.6%) | 5059 (82.6%) | 49825 (80.9%) |
| **Comorbidities** |  |  |  |  |  |  |  |  |  |  |  |  |  |
| Diabetes, n (%) | 7644 (14.5%) | 1009 (11.5%) | 950 (15.4%) | 911 (14.1%) | 851 (14.4%) | 850 (13.9%) | 869 (13.1%) | 811 (14.3%) | 889 (14.4%) | 874 (14.2%) | 831 (13.4%) | 817 (13.3%) | 8653 (14.1%) |
| Dyslipidemia, n (%) | 19453 (36.9%) | 2962 (33.6%) | 2205 (35.8%) | 2323 (36%) | 2194 (37.1%) | 2222 (36.4%) | 2390 (36%) | 2032 (35.9%) | 2221 (36%) | 2275 (37.1%) | 2308 (37.4%) | 2245 (36.6%) | 22415 (36.4%) |
| Hypertension, n (%) | 27971 (53%) | 4896 (55.6%) | 3186 (51.7%) | 3332 (51.7%) | 3176 (53.7%) | 3119 (51.1%) | 3501 (52.7%) | 3089 (54.5%) | 3353 (54.3%) | 3328 (54.2%) | 3387 (54.8%) | 3396 (55.4%) | 32867 (53.4%) |
| Chronic Obstructive Pulmonary Disease, n (%) | 1220 (2.3%) | 185 (2.1%) | 145 (2.4%) | 154 (2.4%) | 159 (2.7%) | 142 (2.3%) | 144 (2.2%) | 115 (2%) | 140 (2.3%) | 138 (2.2%) | 139 (2.3%) | 129 (2.1%) | 1405 (2.3%) |
| Chronic Kidney Disease, n (%) | 5955 (11.3%) | 1212 (13.8%) | 629 (10.2%) | 715 (11.1%) | 617 (10.4%) | 710 (11.6%) | 771 (11.6%) | 670 (11.8%) | 758 (12.3%) | 778 (12.7%) | 723 (11.7%) | 796 (13%) | 7167 (11.6%) |
| Rheumatoid Arthritis, n (%) | 2058 (3.9%) | 256 (2.9%) | 224 (3.6%) | 268 (4.2%) | 270 (4.6%) | 286 (4.7%) | 288 (4.3%) | 219 (3.9%) | 238 (3.9%) | 171 (2.8%) | 180 (2.9%) | 170 (2.8%) | 2314 (3.8%) |
| Myocardial Infarction, n (%) | 363 (0.7%) | 48 (0.5%) | 44 (0.7%) | 50 (0.8%) | 48 (0.8%) | 35 (0.6%) | 43 (0.6%) | 40 (0.7%) | 35 (0.6%) | 32 (0.5%) | 43 (0.7%) | 41 (0.7%) | 411 (0.7%) |
| Stroke, n (%) | 3820 (7.2%) | 710 (8.1%) | 474 (7.7%) | 393 (6.1%) | 439 (7.4%) | 423 (6.9%) | 529 (8%) | 424 (7.5%) | 503 (8.1%) | 485 (7.9%) | 417 (6.7%) | 443 (7.2%) | 4530 (7.4%) |
| Atrial Fibrillation, n (%) | 2857 (5.4%) | 446 (5.1%) | 322 (5.2%) | 338 (5.2%) | 331 (5.6%) | 340 (5.6%) | 362 (5.4%) | 289 (5.1%) | 344 (5.6%) | 331 (5.4%) | 338 (5.5%) | 308 (5%) | 3303 (5.4%) |
| Under steroid therapy, n (%) | 1929 (3.7%) | 175 (2%) | 262 (4.3%) | 249 (3.9%) | 214 (3.6%) | 256 (4.2%) | 247 (3.7%) | 188 (3.3%) | 204 (3.3%) | 208 (3.4%) | 144 (2.3%) | 132 (2.2%) | 2104 (3.4%) |
| **Facility characteristics** |  |  |  |  |  |  |  |  |  |  |  |  |  |
| DPC classification, n (%) |  |  |  |  |  |  |  |  |  |  |  |  |  |
| *Non-DPC* | 34012 (64.5%) | 6377 (72.4%) | 3839 (62.3%) | 4145 (64.3%) | 3678 (62.2%) | 3665 (60.1%) | 4132 (62.2%) | 3506 (61.9%) | 3878 (62.8%) | 3994 (65.1%) | 4580 (74.1%) | 4972 (81.1%) | 40389 (65.6%) |
| *University DPC* | 2944 (5.6%) | 212 (2.4%) | 821 (13.3%) | 710 (11%) | 370 (6.3%) | 403 (6.6%) | 122 (1.8%) | 79 (1.4%) | 243 (3.9%) | 183 (3%) | 225 (3.6%) | 0 (0.0%) | 3156 (5.1%) |
| *Designated Hospital DPC* | 4057 (7.7%) | 460 (5.2%) | 473 (7.7%) | 254 (3.9%) | 384 (6.5%) | 426 (7%) | 614 (9.2%) | 616 (10.9%) | 514 (8.3%) | 763 (12.4%) | 347 (5.6%) | 126 (2.1%) | 4517 (7.3%) |
| *Standard Hospital DPC* | 11739 (22.3%) | 1757 (20%) | 1028 (16.7%) | 1341 (20.8%) | 1482 (25.1%) | 1604 (26.3%) | 1780 (26.8%) | 1463 (25.8%) | 1543 (25%) | 1198 (19.5%) | 1027 (16.6%) | 1030 (16.8%) | 13496 (21.9%) |
| missing, n |  |  |  |  |  |  |  |  |  |  |  |  |  |
| No of beds, n (%) |  |  |  |  |  |  |  |  |  |  |  |  |  |
| *<20 beds* | 23832 (45.2%) | 3935 (44.7%) | 2653 (43.1%) | 3136 (48.6%) | 2724 (46.1%) | 2508 (41.1%) | 2864 (43.1%) | 2316 (40.9%) | 2292 (37.1%) | 2713 (44.2%) | 3081 (49.9%) | 3480 (56.8%) | 27767 (45.1%) |
| *20 - <50 beds* | 927 (1.8%) | 242 (2.8%) | 51 (0.8%) | 38 (0.6%) | 97 (1.6%) | 104 (1.7%) | 117 (1.8%) | 67 (1.2%) | 146 (2.4%) | 210 (3.4%) | 256 (4.1%) | 83 (1.4%) | 1169 (1.9%) |
| *50 - <100 beds* | 3661 (6.9%) | 1047 (11.9%) | 265 (4.3%) | 287 (4.5%) | 340 (5.7%) | 459 (7.5%) | 580 (8.7%) | 456 (8.1%) | 562 (9.1%) | 407 (6.6%) | 607 (9.8%) | 745 (12.2%) | 4708 (7.6%) |
| *100 - <150 beds* | 2566 (4.9%) | 840 (9.5%) | 236 (3.8%) | 232 (3.6%) | 319 (5.4%) | 304 (5%) | 284 (4.3%) | 355 (6.3%) | 364 (5.9%) | 353 (5.8%) | 437 (7.1%) | 522 (8.5%) | 3406 (5.5%) |
| *150 - <200 beds* | 3220 (6.1%) | 690 (7.8%) | 202 (3.3%) | 409 (6.3%) | 270 (4.6%) | 268 (4.4%) | 524 (7.9%) | 451 (8%) | 494 (8%) | 503 (8.2%) | 483 (7.8%) | 306 (5%) | 3910 (6.3%) |
| *200 - <300 beds* | 2918 (5.5%) | 520 (5.9%) | 280 (4.5%) | 243 (3.8%) | 282 (4.8%) | 418 (6.9%) | 445 (6.7%) | 461 (8.1%) | 560 (9.1%) | 301 (4.9%) | 307 (5%) | 141 (2.3%) | 3438 (5.6%) |
| *300 - <400 beds* | 4143 (7.9%) | 543 (6.2%) | 712 (11.6%) | 449 (7%) | 457 (7.7%) | 578 (9.5%) | 537 (8.1%) | 517 (9.1%) | 666 (10.8%) | 420 (6.8%) | 234 (3.8%) | 116 (1.9%) | 4686 (7.6%) |
| *400 - <500 beds* | 2407 (4.6%) | 251 (2.9%) | 136 (2.2%) | 324 (5%) | 318 (5.4%) | 365 (6%) | 346 (5.2%) | 346 (6.1%) | 150 (2.4%) | 167 (2.7%) | 195 (3.2%) | 311 (5.1%) | 2658 (4.3%) |
| *500 - <600 beds* | 2417 (4.6%) | 180 (2%) | 240 (3.9%) | 276 (4.3%) | 331 (5.6%) | 136 (2.2%) | 281 (4.2%) | 260 (4.6%) | 428 (6.9%) | 410 (6.7%) | 0 (0.0%) | 235 (3.8%) | 2597 (4.2%) |
| 600 - <700 beds | 2365 (4.5%) | 272 (3.1%) | 308 (5%) | 299<br>(4.6%) | 327<br>(5.5%) | 340<br>(5.6%) | 302<br>(4.5%) | 221<br>(3.9%) | 244<br>(4%) | 398<br>(6.5%) | 198<br>(3.2%) | 0<br>(0.0%) | 2637<br>(4.3%) |
| 700 - <800 beds | 606 (1.1%) | 133 (1.5%) | 117 (1.9%) | 77<br>(1.2%) | 21<br>(0.4%) | 2<br>(0.03%) | 27<br>(0.4%) | 27<br>(0.5%) | 23<br>(0.4%) | 51<br>(0.8%) | 225<br>(3.6%) | 169<br>(2.8%) | 739<br>(1.2%) |
| 800 - <900 beds | 1809 (3.4%) | 89 (1%) | 259 (4.2%) | 285<br>(4.4%) | 216<br>(3.7%) | 398<br>(6.5%) | 248<br>(3.7%) | 72<br>(1.3%) | 97<br>(1.6%) | 147<br>(2.4%) | 156<br>(2.5%) | 20<br>(0.3%) | 1898<br>(3.1%) |
| 900 beds or over | 1881 (3.6%) | 64 (0.7%) | 702<br>(11.4%) | 395<br>(6.1%) | 212<br>(3.6%) | 218<br>(3.6%) | 93<br>(1.4%) | 115<br>(2%) | 152<br>(2.5%) | 58<br>(0.9%) | 0<br>(0.0%) | 0<br>(0.0%) | 1945<br>(3.2%) |
| Individual Duplicate Counts |  |  |  |  |  |  |  |  |  |  |  |  |  |
| None | 52552 (99.6%) | 8774 (99.6%) | 6140<br>(99.7%) | 6429<br>(99.7%) | 5891<br>(99.6%) | 6078<br>(99.7%) | 6628<br>(99.7%) | 5642<br>(99.6%) | 6154<br>(99.6%) | 6114<br>(99.6%) | 6157<br>(99.6%) | 6093<br>(99.4%) | 61326<br>(99.6%) |
| Once | 200 (0.4%) | 32 (0.4%) | 21 (0.3%) | 21<br>(0.3%) | 23<br>(0.4%) | 20<br>(0.3%) | 20<br>(0.3%) | 22<br>(0.4%) | 24<br>(0.4%) | 24<br>(0.4%) | 22<br>(0.4%) | 35<br>(0.6%) | 232<br>(0.4%) |

Means (SD) or medians [IQR] are presented for continuous data
DPC: Diagnosis Procedure Combination

Patients who were prescribed romosozumab were older than those receiving bisphosphonates (80.5 vs. 78.3 years). For both drugs, there was a majority of females, comprising 80.2% and 85.3% of bisphosphonate and romosozumab users, respectively. The prevalence of diabetes, dyslipidemia, and hypertension among bisphosphonate and romosozumab users was 14.5% and 11.5%, 36.9% and 33.6%, and 53% and 55.6%, respectively. There were also imbalances in the proportions of other complications such as COPD, CKD, rheumatoid arthritis, myocardial infarction within 1 year, stroke within one year, AF, and steroid therapy.

Facility characteristics disparities were even more evident between bisphosphonate and romosozumab users. Romosozumab users were more likely to belong to non-DPC facilities (72.4% vs. 64.5%), to belong to hospitals with a lower number of beds (≥ 20 to < 300 beds), and less likely to belong to hospitals with a greater number of beds ( ≥ 300 beds).

When stratified by deciles based on the instrumental variable of facility-level prescribing preferences, some imbalances in patient characteristics related to CVD were less apparent than those by medications, namely sex, diabetes, dyslipidemia, myocardial infarction, stroke, and AF. Conversely, imbalances in facility-level prescribing preferences were more pronounced than those of medications.

### Incidence of cardiovascular events and secondary outcomes

During the 1-year observation period, the incidence of CVD events among romosozumab users was 7.98 per 100 person-years, while in bisphosphonate users it was 7.15 per 100 person-years (Table 2). The 1-year unadjusted IRR for CVD events between romosozumab and bisphosphonate was 1.12 [95% CI: 1.03-1.21]. During the 2-year observation period, the corresponding incidence of CVD events and unadjusted IRR were 4.60 and 3.97 per 100 patient-years (eTable 6) and 1.16 (95% CI: 1.07-1.26), respectively.

**Table 2.** Incidence rates of the outcome events at one-year.

| *Subjects* (n=61,442) | During one-year |  |  | *IRR* |
| --- | --- | --- | --- | --- |
| *Observations* (n=61,558) | *events* | *person-year* | *incidence rate* |  |
| Cardiovascular Disease |  |  |  |  |
| Bisphosphonate | 3564 | 49867.3 | 7.15 | Ref. |
| Romosozumab | 661 | 8284.0 | 7.98 | 1.12 (1.03, 1.21) |
| Total | 4225 | 58151.3 | 7.27 |  |
| Myocardial Infarction |  |  |  |  |
| Bisphosphonate | 355 | 51877.7 | 0.68 | Ref. |
| Romosozumab | 42 | 8662.5 | 0.49 | 0.71 (0.51, 0.98) |
| Total | 397 | 60540.2 | 0.66 |  |
| Cerebral Infarction |  |  |  |  |
| Bisphosphonate | 2937 | 50260.4 | 5.84 | Ref. |
| Romosozumab | 557 | 8345.2 | 6.67 | 1.14 (1.04, 1.25) |
| Total | 3494 | 58605.6 | 5.96 |  |
| Cerebral Hemorrhage |  |  |  |  |
| Bisphosphonate | 418 | 51841.9 | 0.81 | Ref. |
| Romosozumab | 83 | 8635.2 | 0.96 | 1.19 (0.94, 1.51) |
| Total | 501 | 60477.0 | 0.83 |  |

Incidence rate ratios were estimated by unadjusted Poisson regression models.
IRR: incidence rate ratio

During the 1-year observation period, the incidence of MI, CH, and CI was 0.49, 6.67 and 0.96 per 100 person-years for romosozumab users compared to 0.68, 5.84, and 0.81 per 100 person-years for bisphosphonate users, respectively. During the 2-year observation period, the corresponding values were 0.30, 3.86, and 0.54 per 100 person-years for romosozumab users, compared to 0.40, 3.23, and 0.44 per 100 person-years for bisphosphonate users.

### Comparative cardiovascular safety of romosozumab versus bisphosphonates using the instrumental variable approach

There was a monotonic increase in the likelihood of new prescriptions of romosozumab with increasing deciles of the facility-level proportions of romosozumab prescriptions within the prior 90 days (<u>eTable 7</u>). The goodness of fit of the logistic regression model corresponding to this model was greater than that of the logistic regression model that did not include the decile of the facility-level proportions of romosozumab prescriptions. (pseudo R^2^: 0.134 vs. 0.055).

There was insufficient evidence that a new prescription of romosozumab was associated with increased CVD risk over 1 year compared to bisphosphonates [hazard ratio (HR): 1.09 (95% CI: 0.79-1.76)] (Table 3). The magnitude and precision of the association with CVD over 2 years was similar [HR: 1.00 (95% CI: 0.75-1.61)] (<u>eTable 8)</u>.

**Table 3.** Instrumental variable analyses on the association between new romosozumab prescription versus bisphosphonates and the outcomes.

|  | **During one-year** | **During two-years** |
| --- | --- | --- |
|  | HR (95%CI) | HR (95%CI) |
| Cardiovascular Disease | 1.09 (0.79, 1.76) | 1.00 (0.75, 1.61) |
| Myocardial Infarction | 0.26 (0.07, 1.07) | 0.29 (0.09, 1.21) |
| Cerebral Infarction | 1.21 (0.84, 1.96) | 1.11 (0.79, 1.77) |
| Cerebral Hemorrhage | 1.46 (0.35, 3.00) | 1.52 (0.35, 2.74) |

HR: hazard ratio
Estimated by two-stage residual inclusion method.
In the first stage, new romosozumab prescription was regressed on facility-level proportion of romosozumab prescription using a mixed-effects logistic regression model.
In the second stage, the associations between new romosozumab prescription and the primary and secondary outcomes were estimated by Cox proportional hazards models that included the residual estimated from the first-stage model.

Similarly, there was insufficient evidence to support that a new prescription of romosozumab was associated with excess MI, CI, or CH incidence over 1 and 2 years compared to bisphosphonates [MI: HRs: 0.26 (95% CI: 0.07, 1.07) and 0.29 (95% CI: 0.09, 1.21; CI: HRs: 1.21 (95% CI: 0.84, 1.96) and 1.11 (95% CI: 0.79, 1.77); CH: HRs: 1.46 (95% CI: 0.35, 3.00) and 1.52 (95% CI: 0.35, 2.74), respectively] (eTables 9-11).

## Discussion

Our findings reinforce the lack of clear evidence of a romosozumab-related increase in CVD risk, compared to bisphosphonates among patients with osteoporosis or fragility fractures. as we compared adverse events on a scale beyond the number of subjects treated with romosozumab in previous clinical trials (n=8790). The study findings therefore do not support avoiding romosozumab prescriptions for patients with pre-existing CVD; however, further large-scale pharmacovigilance studies remain necessary.

In the ARCH international phase III trial involving postmenopausal females with a history of fragility fractures, romosozumab (n=2046) was associated with increased serious adverse cardiovascular events at 1 year compared to alendronate.^7^ Similarly, in a placebo-controlled trial involving males with a history of fragility fractures, romosozumab (n=163) was associated with an increase in the incidence of major adverse cardiac events (MACEs).^8^ In contrast, the FRAME trial did not suggest an increased incidence of MACEs associated with romosozumab (n=3589) in postmenopausal women with osteoporosis^32^ A recent meta-analysis including the aforementioned randomized trials comparing romosozumab to placebo or other control treatments also showed no increased risk of serious adverse cardiovascular events or cardiovascular death associated with romosozumab.^33^

Several postmarketing pharmacovigilance studies have suggested a potential increase in CVD events with romosozumab use; however, a lack of true controls (i.e., no CVD events) and confounding factors, plus the presence of reporting bias, make accurate assessment of the relative risk from romosozumab difficult. A database reported to the United States Food and Drug Administration adverse event reporting system revealed a relatively high number of serious CVD events among romosozumab users (n=1995).^9^ Similarly, a study using the Japanese adverse event reporting database indicated a relatively high number of cardiac and cerebrovascular events among romosozumab users (n=859) compared to other osteoporosis medications.^10^ In contrast, the present study excludes selection bias by restricting the subjects to new users^15^ and addresses confounding by indication through comparison of the presence or absence of CVD with the active comparator.^34^ The latter is further minimized by instrumental variable analyses.

Basic research findings also reinforce the debate concerning the association between romosozumab and CVD. The suppression of sclerostin by romosozumab has been suggested to promote vascular calcification,^35,36^ whereas it increases bone mass and strength of cortical and trabecular bone in the skeletal system by neutralizing osteogenesis inhibition.^37–39^ However, basic studies using cynomolgus monkeys, rats, and atherosclerosis-prone mice, sponsored by romosozumab’s manufacturing and marketing companies, did not find signs of promotion of vascular calcification or associated CVD.^40^

The clinical implication of this study includes providing physicians with reassurance on the safety of romosozumab regarding CVD incidence in the management of patients with osteoporosis, as no definitive evidence of increased risk compared to bisphosphonates was detected. Nevertheless, the need for further research regarding the safety of romosozumab remains, since a safety warning was issued by the Japanese Pharmaceuticals and Medical Devices Agency in 2019.

This study had several strengths. First, we included a larger number of romosozumab users than the combined number of romosozumab users in previous clinical trials and observational studies, thereby improving the detection of CVD. Second, we used an active comparator and new-user design to combat biases inherent to observational studies.^15,34^ The new-user design also eliminated selection bias as a result of including patients who are lost to follow-up due to early events of interest or other side effects following osteoporosis drug prescriptions. Third, we addressed unmeasured confounders by using the instrumental variable of facility prescription preferences.^41^ Finally, the use of the DeSC database, which has been validated to be representative of a broader Japanese population,^13^ ensures the generalizability of our study findings.

There are several limitations to the present study. First, due to the use of claims data, medication use documentation, outcome incidence, and the presence or absence of comorbidities may not be completely accurate. The actual administration of the medications and adherence also cannot be ascertained. However, at least the duration of exposure to osteoporosis medications was appropriately defined through the implementation of the refill-gap method, which is standard in pharmacoepidemiologic studies, and through the specification of a clinically plausible grace period.^14^ To account for imprecisions in the diagnostic terms, the definitions of outcome variables and comorbidity variables in this study were defined based on the findings of previous validation studies. For example, the definition of CVD based on a combination of disease/procedure codes, medications, as well as testing, guarantees a high positive predictive value for true CVD events.^17,18^ Second, it is not clear whether our findings can be extrapolated to other racial groups with different CVD risk profiles as the study population was Japanese. Third, the maximum follow-up period of 2 years is relatively short considering the latency period of CVDs. However, we believe it is a reasonable exposure period given that romosozumab is in principle prescribed for 1 year and that the subsequent legacy effect is unknown. Finally, it should be noted that the failure to demonstrate the greater CVD risk of romosozumab does not necessarily mean a lack of difference in CVD incidence. The findings of this study should be interpreted as romosozumab having a potential effect on CVD incidence from a 21% reduction to a 76% increase over 1 year compared to bisphosphonates.^42^

In conclusion, this large pharmacoepidemiological study failed to demonstrate definitive evidence that romosozumab increases CVD risk compared to bisphosphonates. This finding is crucial for the osteoporosis treatment decision-making and challenges clinicians’ undue concern about the safety of romosozumab regarding CVD risk. Our findings should be validated in future studies with longer-term follow-up among different racial groups and larger populations.

## Data Availability

All data produced in the present work are contained in the manuscript

## Author Contributions

Drs Tominaga, Taguri, and Kurita had full access to all of the data in the study and take responsibility for the integrity of the data and the accuracy of the data analysis. *Concept and design*: Tominaga, Ikenoue, Taguri, Kurita. *Acquisition, analysis, or interpretation of data*: All authors. *Drafting of the manuscript*: Tominaga, Kurita. *Critical revision of the manuscript for important intellectual content*: All authors. *Statistical analysis*: Ishii, Okuda, Taguri, Kurita. *Obtained funding*: None. *Administrative, technical, or material support*: Ikenoue, Shimizu, Kurita. *Supervision*: Taguri, Kurita.

## Data Sharing Statement

The dataset analyzed in this paper is available from the corresponding author on reasonable request.

## Additional Contributions

We would like to express our sincere gratitude to Dr. Takashi Igarashi from the Department of Cardiovascular Surgery at Fukushima Medical University, Dr. Takuya Maeda from the Department of Neurosurgery at Fukushima Medical University, and Dr. Susumu Kobayashi from the Department of Neurology from the Kikaijima Tokushukai Hospital, for their invaluable guidance and collaboration in defining the outcomes. Their clinical expertise has greatly contributed to this study.

