## Supplementary Files for "Comparative cardiovascular safety of romosozumab versus bisphosphonates in Japanese patients with osteoporosis A new-user, active comparator design with instrumental variable analyses"

**eTable 1. List of the codes for eligibility criteria**

**eTable 2. Bisphosphonates listed as indicated for osteoporosis in Japan**

**eTable 3. Codes for outcome definition**

**eTable 4. Definition and codes for covariates**

**eTable 5. Glucocorticoids and WHO-ATC codes**

**eTable 6. Incidence rates of the outcome events at 2 years**

**eTable 7. Association of romosozumab prescription with facility-level preference and patient and facility characteristics**

**eTable 8. IV analyses on the association between romosozumab and cardiovascular events**

**eTable 9. IV analyses on the association between romosozumab and myocardial infarction**

**eTable 10. IV analyses on the association between romosozumab and cerebral infarction**

**eTable 11. IV analyses on the association between romosozumab and cerebral hemorrhage**

**eFigure 1. Directed acyclic graph of the present study**

**eMethods 1. Definition of outcomes of the present study**

**eMethods 2. The detailed descriptions of the reason why observations with a facility-level prescription proportion of zero were excluded**

**eTable 1. List of the codes for eligibility criteria**

| **Category** | **Details** | **Codes** | |
| --- | --- | --- | --- |
| Diagnosis of Osteoprosis | ICD-10 | M80, M81 |  |
| Anti-osteporotic drugs | WHO-ATC | M05 | Drugs for Treatment of Bone Diseases |
|  |  | A11CC | Vitamin D and analogues |
|  |  | A12A | Calcium |
|  |  | A12CD | Fluoride |
|  |  | A14AA04 | Metenolone |
|  |  | B02BA | Vitamin K |
|  |  | G03C | Estrogens |
|  |  | G03F | Progestogens and Estrogens in Combination |
|  |  | G03XC | Selective estrogen receptor modulators |
|  |  | H05AA02 | Teriparatide |
|  |  | H05BA | Calcitonin preparations |
| Frailty fracture location |  |  |  |
| Femur | ICD-10 | S72.0, S72.1, S72.2 |  |
| Distal Radius | ICD-10 | S52.5 |  |
| Proximal Humerus | ICD-10 | S42.2 |  |
| Thoracic Vertebra | ICD-10 | S22.0 |  |
| Lumbar Vertebra | ICD-10 | S32.0 |  |

ICD-10: the International Classification of Diseases, 10th revision (ICD-10), WHO-ATC: World Health Organization Anatomical Therapeutic Chemical

**eTable 2. Bisphosphonates listed as indicated for osteoporosis in Japan**

| **Drug name** | **WHO-ATC** | **MHLW Code** |
| --- | --- | --- |
| Etidronate | M05BA01 |  |
| Alendronate | M05BA04 |  |
| Risedronate | M05BA07 |  |
| Minodronate | - | 3999026^a^ |
| Ibandronate | M05BA06 |  |
| Zoledronate | M05BA08 | 3999423A4027^b^ |

^a^ First seven digits of the MHLW code

^b^ Zometa/Zoledronic acid is not indicated for osteoporosis in Japan. Only Reclast (identified with the YJ code and the individual drug code 3999423A4027 ) is indicated for osteoporosis

WHO-ATC: World Health Organization Anatomical Therapeutic Chemical; MHLW: Ministry of Health, Labour and Welfare

**eTable 3. Codes for outcome definition**

|  | **Outcome** |  | **Description** | **Codes** |
| --- | --- | --- | --- | --- |
|  | **CVD (Composite variable)** | **Definition** | **Myocardial Infarction [OR] Stroke** |  |
|  | **Myocardial Infarction (MI)** | **Definition** | **Diagnosis [AND] (Medical Procedures [OR] Drugs) [AND] Hospitalization.** |  |
|  |  | Diagnosis | ICD-10 Code | I20, I21, I22, I23, I24 |
|  |  | Medical Procedure | Surgery [OR] Rehabilitation | 150374910, 150375010, 150375110, 150260350, 150284310, 150359310, 150375210, 150375310, 150375410, 160107550, 150318310, 150145710, 150145810, 150145910, 150146010, 150302770, 150318410, 150318510, 150143110, 150318610, 150318810, 150319110, 150319410, 150318910, 150319210, 150319510, 180027410, 180027510 |
|  |  | Drugs for Myocardial Infarction | WHO-ATC code | Urokinase (B01AD04), Alteplase (ATC code B01AD02), Monteplase (first seven digits of the MHLW code 3959407) |
|  | **Stroke** | **Definition** | **Diagnosis [AND] (Medical Procedures [OR] Drugs) [AND] Hospitalization [AND] Imaging** |  |
|  |  | Diagnosis | ICD-10 Code | I60, I61, I63, I64, G459 |
|  | **eTable 3. Codes for outcome definition (continued)** | | | |
|  |  | Medical Procedure | Surgery [OR] Rehabilitation | 150066210, 150067110, 150411910, 150067210, 150067410, 150069610, 150069850, 150069950, 150335710, 150372310, 150243410, 150243510, 150243610, 150243710, 150243810, 150243910, 150254910, 150344410, 150355410, 150273510, 150301110, 150301210, 150372510, 150380850, 180027410, 180027510, 180027610, 180027710, 180030810, 180044310, 180044410, 180044510, 180050330, 180050430, 180050530, 180050630, 180050830, 180051030 |
|  |  | Drugs for Cerebral Infarction | WHO-ATC code | Urokinase (B01AD04), Alteplase (B01AD02), Edaravone (N07XX14), Argatroban (B01AE03), Ozagrel Sodium (first seven digits of the MHLW code 3999411), Heparin (B01AB01, B01AB12) |
|  |  | Imaging | (CT [OR] MRI) within 30 days of diagnosis. | 170011710, 170011810, 170028610, 170033410, 170034910, 170038710, 170038810, 170038910, 170039010, 170039110, 170040210, 170040310, 170040410, 170040510, 170040610, 170040710, 170040810, 170040910, 170041010, 170041110, 170033510, 170035010, 170041410, 170041510, 170020110, 170041610, 170015210, 170041710 |
|  | **Cerebral Infarction** | **Definition** | **Diagnosis [AND] Drugs [AND] Hospitalization [AND] Imaging** |  |
|  |  | Diagnosis | ICD-10 Code | I63, I64, G459 |
|  |  | Drugs for Cerebral Infarction | WHO-ATC code | Urokinase (B01AD04), Alteplase (B01AD02), Edaravone (N07XX14), Argatroban (B01AE03), Ozagrel Sodium (first seven digits of the MHLW code: 3999411), Heparin (B01AB01, B01AB12) |
| **eTable 3. Codes for outcome definition (continued)** | | | | |
|  |  | Imaging | (CT [OR] MRI) within 30 days of diagnosis. | 170011710, 170011810, 170028610, 170033410, 170034910, 170038710, 170038810, 170038910, 170039010, 170039110, 170040210, 170040310, 170040410, 170040510, 170040610, 170040710, 170040810, 170040910, 170041010, 170041110, 170033510, 170035010, 170041410, 170041510, 170020110, 170041610, 170015210, 170041710 |
|  | **Cerebral Hemorrhage** | **Definition** | **Diagnosis [AND] (Medical Procedures [OR] Drugs) [AND] Hospitalization [AND] Imaging** |  |
|  |  | Diagnosis | ICD-10 Code | I60, I61 |
|  |  | Medical Procedure | Surgery [OR] Rehabilitation | 150066210, 150067110, 150411910, 150067210, 150067410, 150069610, 150069850, 150069950, 150335710, 150372310, 150243410, 150243510, 150243610, 150243710, 150243810, 150243910, 150254910, 150344410, 150355410, 150273510, 150301110, 150301210, 150372510, 150380850, 180027410, 180027510, 180027610, 180027710, 180030810, 180044310, 180044410, 180044510, 180050330, 180050430, 180050530, 180050630, 180050830, 180051030 |
| **eTable 3. Codes for outcome definition (continued)** | | | | |
|  |  | Imaging | (CT [OR] MRI) within 30 days of diagnosis. | 170011710, 170011810, 170028610, 170033410, 170034910, 170038710, 170038810, 170038910, 170039010, 170039110, 170040210, 170040310, 170040410, 170040510, 170040610, 170040710, 170040810, 170040910, 170041010, 170041110, 170033510, 170035010, 170041410, 170041510, 170020110, 170041610, 170015210, 170041710 |

ICD-10: the International Classification of Diseases, 10th revision (ICD-10), WHO-ATC: World Health Organization Anatomical Therapeutic Chemical

**eTable 4. Definition and codes for covariates**

| **Covariate** |  | **Description** | **Codes** |
| --- | --- | --- | --- |
| **Diabetes** | **Definition** | **Diagnosis [AND] Drugs** |  |
|  | Diagnosis | ICD-10 Code | E10, E11, E12, E13, E14 |
|  | Drugs for diabetes | WHO-ATC code | A10AB, A10AC, A10AD, A10AE, A10AF, A10BA, A10BB, A10BC, A10BD, A10BF, A10BG, A10BH, A10BJ, A10BK, A10BX, A10XA |
| **Hypertension** | **Definition** | **Diagnosis [AND] Drugs** |  |
|  | Diagnosis | ICD-10 Code | I10, I11, I12, I13, I15 |
|  | Drugs for Hypertension | WHO-ATC code | C02C, C03A, C07A, C08C, C08D, C09A, C09B, C09C, C09D, C09X, C10BX03, C10BX07, C10BX09, C10BX10, C10BX11, C10BX12, C10BX13, C10BX14, C10BX15, C10BX16, C10BX17, C10BX18 |
| **Dyslipidemia** | **Definition** | **Diagnosis [AND] Drugs** |  |
|  | Diagnosis | ICD-10 Code | E78 |
|  | Drugs for Dyslipidemia | WHO-ATC code | A11HA03, C10AA01, C10AA02, C10AA03, C10AA04, C10AA05, C10AA07, C10AB01, C10AB02, C10AB03, C10AB05, C10AB12, C10AC01, C10AD02, C10AX02, C10AX06, C10AX09, C10AX12, C10AX13, C10AX14, C10BA02, C10BA05, C10BX03, V03AE06, Nicomol(first seven digits of the MHLW code: 2189004) |
| **Chronic obstructive pulmonary disease (COPD)** | **Definition** | **Diagnosis [AND] Drugs** |  |
|  | Diagnosis | ICD-10 Code | J41, J43, J44 |
|  | Drugs for COPD | WHO-ATC code | R03AC, R03AK, R03AL, R03BB |
| **eTable 4. Definition and codes for covariates (continued)** | | | |
| **Chronic kidney disease (CKD)** | **Definition** | **Diagnosis** |  |
|  | Diagnosis | ICD-10 Code | N05.5, N05.6, N05.7, N18, N19, N25.0, Z49.0, Z49.1, Z49.2, Z94.0, Z99.2 |
| **Rheumatoid arthritis (RA)** | **Definition** | **Diagnosis [AND] Drugs** |  |
|  | Diagnosis | ICD-10 Code | M05, M06 |
|  | Drugs for RA | WHO-ATC code | A07EC01, D11AH01, L04AA13, L04AA24, L04AA29, L04AA37, L04AA44, L04AA45, L04AA49, L04AB01, L04AB02, L04AB04, L04AB05, L04AB06, L04AC07, L04AC14, L04AX01, L04AX03, M01CB01, M01CB03, M01CC01, M01CC02, Actarit (first seven digits of the MHLW code: 1149031), Iguratimod (first seven digits of the MHLW code: 3999031), Mizoribine (first seven digits of the MHLW code: 3999002) |
| **Atrial fibrillation (Af)** | **Definition** | **Diagnosis [AND] Drugs** |  |
|  | Diagnosis | ICD-10 Code | I48 |
|  | Drugs for Af | WHO-ATC code | B01AA, B01AE, B01AF |

ICD-10: the International Classification of Diseases, 10th revision (ICD-10), WHO-ATC: World Health Organization Anatomical Therapeutic Chemical

**eTable 5. Glucocorticoids and WHO-ATC Codes**

| **Drug** | **WHO-ATC Code** |
| --- | --- |
| Cortisone Acetate | H02AB10, S01BA03 |
| Hydrocortisone | A01AC03, A07EA02, C05AA01, D07AA02, D07XA01, H02AB09, S01BA02, S01CB03, S02BA01 |
| Prednisolone | A01AC04, A07EA01, C05AA04, D07AA03, D07XA02, H02AB06, R01AD02, S01BA04, S01CB02, S02BA03, S03BA02 |
| Methylprednisolone | D07AA01, D10AA02, H02AB04 |
| Triamcinolone | A01AC01, C05AA12, D07AB09, D07XB02, H02AB08, R01AD11, R03BA06, S01BA05 |
| Dexamethasone | H02AB02 |
| Betamethasone | A07EA04, C05AA05, D07AC01, D07XC01, H02AB01, R01AD06, R03BA04, S01BA06, S01CB04, S02BA07, S03BA03 |
| Ciclesonide | R01AD13, R03BA08 |
| Fluticasone Furoate | R03BA05, R03BA09 |
| Fluticasone Propionate | R03BA05 |
| Beclometasone Dipropionate | R03BA01 |
| Mometasone Furoate | R03BA07 |
| Budesonide | R01AD05, R03BA02 |
| Flurandrenolide | D07AC07 |

WHO-ATC: World Health Organization Anatomical Therapeutic Chemical

**eTable 6. Incidence rates of the outcome events at 2 years**

| *Subjects* (n=61,442) | **During 2 years** | |  |  |
| --- | --- | --- | --- | --- |
| *Observations* (n=61,558) | *Events* | *Person-year* | *Incidence rate* | *IRR* |
| **Cardiovascular Disease** |  |  |  |  |
| Bisphosphonate | 3869 | 97581.0 | 3.97 | Ref. |
| Romosozumab | 742 | 16121.9 | 4.60 | 1.16 (1.07, 1.26) |
| Total | 4611 | 113703.0 | 4.06 |  |
| **Myocardial Infarction** |  |  |  |  |
| Bisphosphonate | 406 | 102805.2 | 0.40 | Ref. |
| Romosozumab | 51 | 17137.6 | 0.30 | 0.75 (0.56, 1.01) |
| Total | 457 | 119942.9 | 0.38 |  |
| **Cerebral Infarction** |  |  |  |  |
| Bisphosphonate | 3181 | 98595.2 | 3.23 | Ref. |
| Romosozumab | 628 | 16286.8 | 3.86 | 1.20 (1.10, 1.30) |
| Total | 3809 | 114882.0 | 3.32 |  |
| **Cerebral Hemorrhage** |  |  |  |  |
| Bisphosphonate | 452 | 102721.0 | 0.44 | Ref. |
| Romosozumab | 92 | 17070.5 | 0.54 | 1.22 (0.98, 1.53) |
| Total | 544 | 119791.5 | 0.45 |  |

Incidence rate ratios were estimated by unadjusted Poisson regression models.

IRR: incidence rate ratio

**eTable 7. Association of romosozumab prescription with facility-level preference and patient and facility characteristics**

|  | OR (95%CI) | P-value |
| --- | --- | --- |
| Preference decile |  |  |
| *1st* | Ref. |  |
| *2nd* | 1.40 (1.18, 1.65) | < 0.001 |
| *3rd* | 1.88 (1.60, 2.22) | < 0.001 |
| *4th* | 2.28 (1.94, 2.67) | < 0.001 |
| *5th* | 2.70 (2.32, 3.15) | < 0.001 |
| *6th* | 3.63 (3.11, 4.22) | < 0.001 |
| *7th* | 4.50 (3.88, 5.22) | < 0.001 |
| *8th* | 5.47 (4.72, 6.33) | < 0.001 |
| *9th* | 7.22 (6.25, 8.34) | < 0.001 |
| *10th* | 9.42 (8.16, 10.9) | < 0.001 |
| Age, years | 1.03 (1.02, 1.03) | < 0.001 |
| Female sex | 1.32 (1.24, 1.42) | < 0.001 |
| Comorbidities |  |  |
| *Diabetes* | 0.82 (0.76, 0.88) | < 0.001 |
| *Dyslipidemia* | 0.86 (0.81, 0.90) | < 0.001 |
| *Hypertension* | 1.03 (0.98, 1.08) | 0.32 |
| *Chronic Obstructive Pulmonary Disease* | 1.09 (0.93, 1.29) | 0.285 |
| *Chronic Kidney Disease* | 1.23 (1.14, 1.32) | < 0.001 |
| *Rheumatoid Arthritis* | 1.02 (0.88, 1.17) | 0.811 |
| *Myocardial Infarction* | 0.91 (0.66, 1.25) | 0.56 |
| *Stroke* | 1.11 (1.01, 1.21) | 0.026 |
| *Atrial Fibrillation* | 0.88 (0.79, 0.98) | 0.02 |
| *Under steroid therapy* | 0.75 (0.64, 0.89) | 0.001 |
| *DPC classification* |  |  |
| *Non-DPC* | Ref. | < 0.001 |
| *University DPC* | 2.19 (1.70, 2.82) | < 0.001 |
| *Designated Hospital DPC* | 1.84 (1.55, 2.19) | 0.008 |
| *Standard Hospital DPC* | 1.13 (1.03, 1.24) | 0.001 |
| *No of beds* |  |  |
| *<20 beds* | Ref. |  |
| *20 - <50 beds* | 1.28 (1.11, 1.49) | 0.001 |
| *50 - <100 beds* | 1.46 (1.35, 1.59) | < 0.001 |
| *100 - <150 beds* | 1.65 (1.50, 1.81) | < 0.001 |
| *150 - <200 beds* | 1.20 (1.09, 1.32) | < 0.001 |
| *200 - <300 beds* | 1.10 (0.97, 1.24) | 0.141 |
| *300 - <400 beds* | 0.89 (0.79, 1.01) | 0.075 |
| *400 - <500 beds* | 0.55 (0.47, 0.66) | < 0.001 |
| *500 - <600 beds* | 0.37 (0.30, 0.45) | < 0.001 |
| *600 - <700 beds* | 0.61 (0.50, 0.73) | < 0.001 |
| *700 - <800 beds* | 0.66 (0.50, 0.87) | 0.003 |
| *800 - <900 beds* | 0.25 (0.19, 0.33) | < 0.001 |
| *900 beds or over* | 0.23 (0.16, 0.32) | < 0.001 |

New romosozumab prescription was regressed on facility-level proportion of romosozumab prescription using a mixed-effects logistic regression model. The model's effectiveness is measured using the Nagelkerke Pseudo R-squared statistic. For the model that includes only baseline covariates, the Pseudo R-squared value is 0.055. When facility preference is added to the baseline covariates, the Pseudo R-squared value increases to 0.134.

OR: Odds Ratio, DPC: Diagnosis Procedure Combination

**eTable 8. IV analyses on the association between romosozumab and cardiovascular events**

|  | 1 year |  | 2 years |
| --- | --- | --- | --- |
|  | HR (95%CI) |  | HR (95%CI) |
| Romosozumab | 1.09 (0.79, 1.76) |  | 1.00 (0.75, 1.61) |
| Age, years | 1.02 (1.02, 1.03) |  | 1.02 (1.02, 1.03) |
| Female sex | 0.76 (0.70, 0.82) |  | 0.77 (0.72, 0.84) |
| Comorbidities |  |  |  |
| *Diabetes* | 1.17 (1.08, 1.28) |  | 1.15 (1.06, 1.25) |
| *Dyslipidemia* | 1.07 (1.00, 1.14) |  | 1.07 (1.00, 1.14) |
| *Hypertension* | 1.25 (1.18, 1.34) |  | 1.27 (1.19, 1.35) |
| *Chronic Obstructive Pulmonary Disease* | 1.02 (0.84, 1.25) |  | 1.03 (0.85, 1.25) |
| *Chronic Kidney Disease* | 1.08 (0.98, 1.18) |  | 1.11 (1.02, 1.21) |
| *Rheumatoid Arthritis* | 0.97 (0.79, 1.16) |  | 0.94 (0.79, 1.12) |
| *Myocardial Infarction* | 3.31 (2.58, 4.36) |  | 3.25 (2.56, 4.22) |
| *Stroke* | 13.82 (13.01, 14.74) |  | 12.83 (12.11, 13.66) |
| *Atrial Fibrillation* | 1.18 (1.05, 1.32) |  | 1.17 (1.04, 1.30) |
| *Under steroid therapy* | 1.10 (0.91, 1.29) |  | 1.10 (0.93, 1.29) |
| *DPC classification* |  |  |  |
| *Non-DPC* | Ref. |  | Ref. |
| *University DPC* | 1.14 (0.86, 1.49) |  | 1.18 (0.90, 1.55) |
| *Designated Hospital DPC* | 1.03 (0.82, 1.27) |  | 1.04 (0.85, 1.27) |
| *Standard Hospital DPC* | 0.97 (0.85, 1.09) |  | 0.95 (0.85, 1.07) |
| *No of beds* |  |  |  |
| *<20 beds* | Ref. |  | Ref. |
| *20 - <50 beds* | 1.21 (0.97, 1.48) |  | 1.24 (1.00, 1.50) |
| *50 - <100 beds* | 1.06 (0.93, 1.19) |  | 1.07 (0.96, 1.21) |
| *100 - <150 beds* | 0.98 (0.83, 1.13) |  | 1.01 (0.86, 1.16) |
| *150 - <200 beds* | 1.00 (0.86, 1.13) |  | 1.05 (0.92, 1.18) |
| *200 - <300 beds* | 1.08 (0.92, 1.27) |  | 1.11 (0.94, 1.29) |
| *300 - <400 beds* | 1.18 (1.01, 1.35) |  | 1.17 (1.02, 1.33) |
| *400 - <500 beds* | 1.17 (0.96, 1.44) |  | 1.18 (0.98, 1.42) |
| *500 - <600 beds* | 1.05 (0.84, 1.33) |  | 1.06 (0.86, 1.33) |
| *600 - <700 beds* | 0.91 (0.72, 1.16) |  | 0.89 (0.70, 1.14) |
| *700 - <800 beds* | 1.05 (0.71, 1.45) |  | 1.11 (0.77, 1.52) |
| *800 - <900 beds* | 0.95 (0.74, 1.27) |  | 0.93 (0.72, 1.22) |
| *900 beds or over* | 0.90 (0.68, 1.25) |  | 0.86 (0.65, 1.16) |

Estimated by Cox proportional hazards models including the residual estimated in the first-stage model

HR: hazard ratio, DPC: Diagnosis Procedure Combination

**eTable 9. IV analyses on the association between romosozumab and myocardial infarction**

|  | 1 year |  | 2 years |
| --- | --- | --- | --- |
|  | HR (95%CI) |  | HR (95%CI) |
| Romosozumab | 0.26 (0.07, 1.07) |  | 0.29 (0.09, 1.21) |
| Age, years | 1.03 (1.01, 1.04) |  | 1.03 (1.01, 1.04) |
| Female sex | 0.55 (0.43, 0.69) |  | 0.56 (0.45, 0.69) |
| Comorbidities |  |  |  |
| *Diabetes* | 1.84 (1.46, 2.32) |  | 1.62 (1.30, 2.02) |
| *Dyslipidemia* | 1.20 (0.97, 1.51) |  | 1.20 (0.99, 1.49) |
| *Hypertension* | 2.04 (1.61, 2.67) |  | 1.97 (1.59, 2.47) |
| *Chronic Obstructive Pulmonary Disease* | 1.17 (0.59, 1.90) |  | 1.22 (0.67, 1.92) |
| *Chronic Kidney Disease* | 1.12 (0.84, 1.43) |  | 1.27 (0.98, 1.58) |
| *Rheumatoid Arthritis* | 1.30 (0.75, 1.94) |  | 1.30 (0.79, 1.92) |
| *Myocardial Infarction* | 29.02 (21.67, 39.57) |  | 26.9 (20.2, 36.0) |
| *Stroke* | 1.02 (0.70, 1.38) |  | 0.99 (0.70, 1.33) |
| *Atrial Fibrillation* | 1.96 (1.50, 2.60) |  | 1.83 (1.44, 2.41) |
| *Under steroid therapy* | 1.19 (0.71, 1.81) |  | 1.00 (0.59, 1.48) |
| *DPC classification* |  |  |  |
| *Non-DPC* | Ref. |  | Ref. |
| *University DPC* | 1.71 (0.66, 4.12) |  | 2.29 (0.89, 5.09) |
| *Designated Hospital DPC* | 1.76 (0.89, 3.27) |  | 1.98 (1.07, 3.54) |
| *Standard Hospital DPC* | 1.86 (1.20, 2.83) |  | 1.94 (1.25, 2.95) |
| *No of beds* |  |  |  |
| *<20 beds* | Ref. |  | Ref. |
| *20 - <50 beds* | 1.38 (0.49, 2.51) |  | 1.19 (0.43, 2.13) |
| *50 - <100 beds* | 1.04 (0.63, 1.56) |  | 1.07 (0.68, 1.54) |
| *100 - <150 beds* | 0.66 (0.30, 1.09) |  | 0.73 (0.39, 1.14) |
| *150 - <200 beds* | 0.72 (0.38, 1.17) |  | 0.66 (0.34, 1.06) |
| *200 - <300 beds* | 0.69 (0.34, 1.22) |  | 0.65 (0.34, 1.10) |
| *300 - <400 beds* | 1.26 (0.79, 1.91) |  | 1.13 (0.71, 1.72) |
| *400 - <500 beds* | 0.60 (0.30, 1.11) |  | 0.68 (0.36, 1.20) |
| *500 - <600 beds* | 0.92 (0.47, 1.72) |  | 1.00 (0.55, 1.75) |
| *600 - <700 beds* | 0.60 (0.28, 1.19) |  | 0.59 (0.30, 1.12) |
| *700 - <800 beds* | 0.53 (0.10, 1.33) |  | 0.55 (0.16, 1.29) |
| *800 - <900 beds* | 0.53 (0.20, 1.24) |  | 0.49 (0.20, 1.15) |
| *900 beds or over* | 0.53 (0.17, 1.43) |  | 0.46 (0.16, 1.26) |

Estimated by Cox proportional hazards models including the residual estimated in the first-stage model

HR: hazard ratio, DPC: Diagnosis Procedure Combination

**eTable 10. IV analyses on the association between romosozumab and cerebral infarction**

|  | 1 year |  | 2 years |
| --- | --- | --- | --- |
|  | HR (95%CI) |  | HR (95%CI) |
| Romosozumab | 1.21 (0.84, 1.96) |  | 1.11 (0.79, 1.77) |
| Age, years | 1.03 (1.02, 1.03) |  | 1.03 (1.02, 1.03) |
| Female sex | 0.80 (0.73, 0.87) |  | 0.81 (0.74, 0.88) |
| Comorbidities |  |  |  |
| *Diabetes* | 1.15 (1.05, 1.26) |  | 1.14 (1.05, 1.25) |
| *Dyslipidemia* | 1.11 (1.04, 1.19) |  | 1.11 (1.04, 1.19) |
| *Hypertension* | 1.20 (1.12, 1.30) |  | 1.23 (1.14, 1.32) |
| *Chronic Obstructive Pulmonary Disease* | 1.05 (0.84, 1.29) |  | 1.05 (0.84, 1.28) |
| *Chronic Kidney Disease* | 1.11 (1.00, 1.22) |  | 1.13 (1.02, 1.24) |
| *Rheumatoid Arthritis* | 0.96 (0.78, 1.17) |  | 0.92 (0.74, 1.10) |
| *Myocardial Infarction* | 0.94 (0.65, 1.30) |  | 0.9 (0.63, 1.23) |
| *Stroke* | 15.9 (14.9, 17.1) |  | 14.7 (13.8, 15.7) |
| *Atrial Fibrillation* | 1.17 (1.05, 1.32) |  | 1.17 (1.04, 1.31) |
| *Under steroid therapy* | 1.01 (0.82, 1.21) |  | 1.04 (0.85, 1.24) |
| *DPC classification* |  |  |  |
| *Non-DPC* | Ref. |  | Ref. |
| *University DPC* | 0.99 (0.74, 1.34) |  | 1.02 (0.76, 1.34) |
| *Designated Hospital DPC* | 0.85 (0.67, 1.07) |  | 0.87 (0.68, 1.09) |
| *Standard Hospital DPC* | 0.89 (0.79, 1.01) |  | 0.88 (0.78, 0.99) |
| *No of beds* |  |  |  |
| *<20 beds* | Ref. |  | Ref. |
| *20 - <50 beds* | 1.24 (0.97, 1.52) |  | 1.27 (1.00, 1.55) |
| *50 - <100 beds* | 1.01 (0.88, 1.16) |  | 1.03 (0.90, 1.16) |
| *100 - <150 beds* | 0.96 (0.81, 1.12) |  | 0.99 (0.84, 1.14) |
| *150 - <200 beds* | 0.99 (0.85, 1.14) |  | 1.07 (0.92, 1.22) |
| *200 - <300 beds* | 1.17 (0.97, 1.38) |  | 1.18 (0.98, 1.38) |
| *300 - <400 beds* | 1.11 (0.95, 1.30) |  | 1.11 (0.96, 1.29) |
| *400 - <500 beds* | 1.29 (1.03, 1.61) |  | 1.25 (1.01, 1.54) |
| *500 - <600 beds* | 1.11 (0.87, 1.41) |  | 1.08 (0.85, 1.37) |
| *600 - <700 beds* | 0.99 (0.76, 1.29) |  | 0.96 (0.74, 1.26) |
| *700 - <800 beds* | 1.17 (0.77, 1.64) |  | 1.23 (0.83, 1.66) |
| *800 - <900 beds* | 1.19 (0.87, 1.61) |  | 1.15 (0.86, 1.56) |
| *900 beds or over* | 1.04 (0.76, 1.44) |  | 0.98 (0.72, 1.34) |

Estimated by Cox proportional hazards models including the residual estimated in the first-stage model

HR: hazard ratio, DPC: Diagnosis Procedure Combination

**eTable 11. IV analyses on the association between romosozumab and cerebral hemorrhage**

|  | 1 year |  | 2 years |
| --- | --- | --- | --- |
|  | HR (95%CI) |  | HR (95%CI) |
| Romosozumab | 1.46 (0.35, 3.00) |  | 1.52 (0.35, 2.74) |
| Age, years | 0.98 (0.97, 0.99) |  | 0.98 (0.97, 1.00) |
| Women | 0.86 (0.70, 1.10) |  | 0.87 (0.72, 1.11) |
| Comorbidities |  |  |  |
| *Diabetes* | 0.75 (0.55, 0.97) |  | 0.73 (0.55, 0.93) |
| *Dyslipidemia* | 0.73 (0.59, 0.87) |  | 0.73 (0.59, 0.87) |
| *Hypertension* | 1.19 (0.99, 1.45) |  | 1.16 (0.97, 1.41) |
| *Chronic Obstructive Pulmonary Disease* | 0.85 (0.42, 1.33) |  | 0.78 (0.38, 1.23) |
| *Chronic Kidney Disease* | 0.88 (0.65, 1.15) |  | 0.93 (0.71, 1.19) |
| *Rheumatoid Arthritis* | 0.81 (0.40, 1.25) |  | 0.85 (0.46, 1.29) |
| *Myocardial Infarction* | 1.41 (0.44, 2.74) |  | 1.32 (0.41, 2.58) |
| *Stroke* | 16.17 (13.26, 19.67) |  | 15.25 (12.59, 18.56) |
| *Atrial Fibrillation* | 0.84 (0.55, 1.18) |  | 0.83 (0.55, 1.14) |
| *Under steroid therapy* | 1.24 (0.73, 1.84) |  | 1.26 (0.77, 1.80) |
| *DPC classification* |  |  |  |
| *Non-DPC* | Ref. |  | Ref. |
| *University DPC* | 2.17 (1.00, 4.72) |  | 1.97 (0.95, 4.11) |
| *Designated Hospital DPC* | 2.11 (1.28, 3.80) |  | 1.80 (1.09, 3.12) |
| *Standard Hospital DPC* | 1.37 (0.98, 1.89) |  | 1.24 (0.91, 1.70) |
| *No of beds* |  |  |  |
| *<20 beds* | Ref. |  | Ref. |
| *20 - <50 beds* | 1.04 (0.45, 1.83) |  | 1.06 (0.48, 1.86) |
| *50 - <100 beds* | 1.38 (1.00, 1.92) |  | 1.43 (1.07, 1.97) |
| *100 - <150 beds* | 1.24 (0.87, 1.76) |  | 1.33 (0.95, 1.90) |
| *150 - <200 beds* | 0.98 (0.63, 1.43) |  | 1.05 (0.68, 1.48) |
| *200 - <300 beds* | 0.69 (0.39, 1.08) |  | 0.82 (0.49, 1.27) |
| *300 - <400 beds* | 0.93 (0.57, 1.43) |  | 1.00 (0.64, 1.48) |
| *400 - <500 beds* | 0.81 (0.43, 1.27) |  | 1.04 (0.56, 1.64) |
| *500 - <600 beds* | 1.06 (0.58, 1.68) |  | 1.18 (0.65, 1.81) |
| *600 - <700 beds* | 0.47 (0.20, 0.88) |  | 0.56 (0.25, 1.01) |
| *700 - <800 beds* | 1.06 (0.37, 2.30) |  | 1.31 (0.50, 2.72) |
| *800 - <900 beds* | 0.49 (0.20, 0.97) |  | 0.53 (0.22, 1.03) |
| *900 beds or over* | 0.37 (0.12, 0.84) |  | 0.43 (0.15, 0.91) |

Estimated by Cox proportional hazards models including the residual estimated in the first-stage model

HR: hazard ratio, DPC: Diagnosis Procedure Combination

**eFigure 1. Directed acyclic graph of the present study**

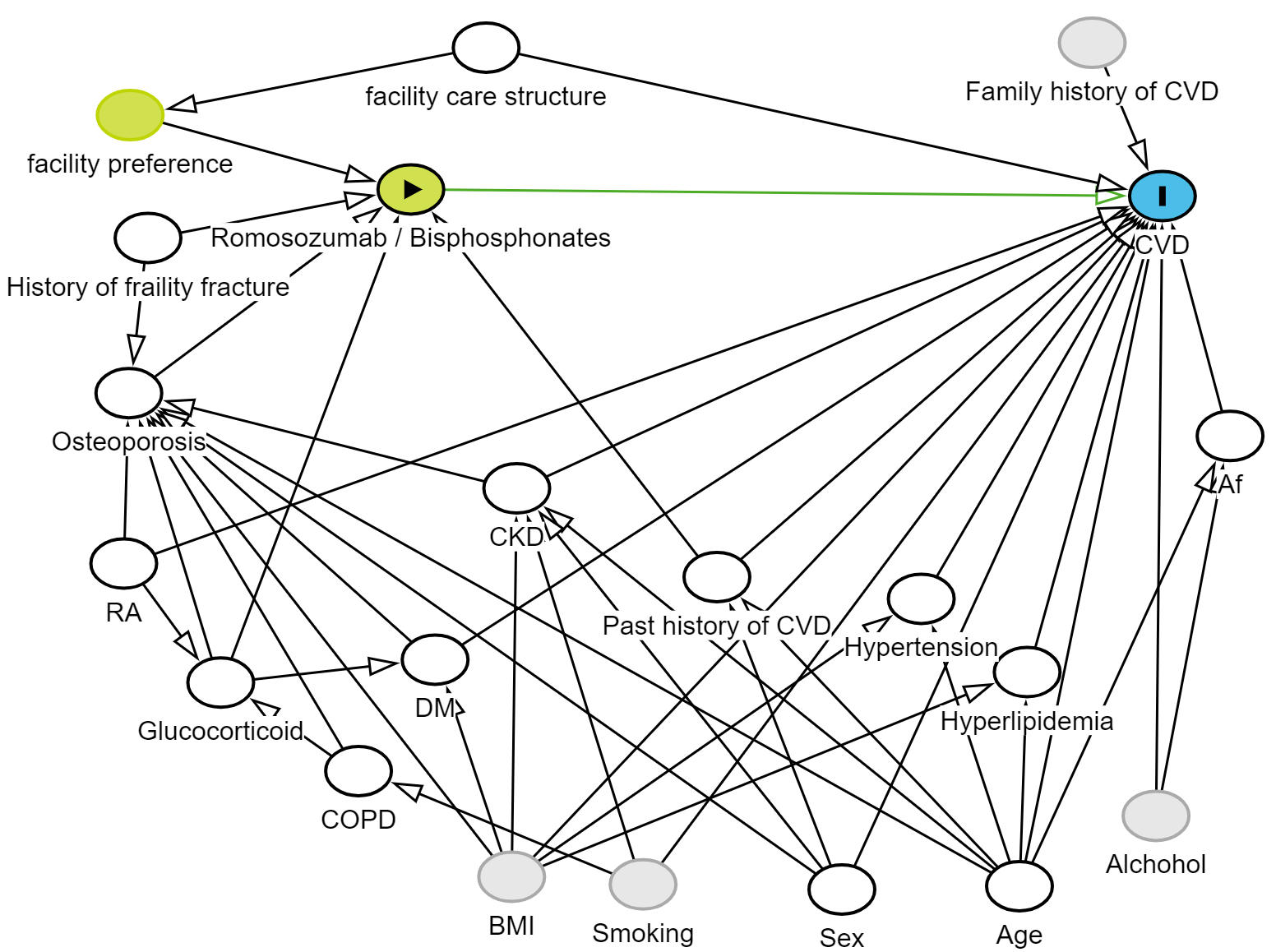
Causal relationship assumptions are shown in these schemas. The yellow-green ovals indicate osteoporosis medication (“exposure”), and the sky-blue ovals indicate cardiovascular disease (“outcome”). The white ovals are ancestors of both the exposure and outcome and represent potential confounders, while the plain green and grey ovals are ancestors of the exposure and unobserved variables, respectively. The green lines, which show potential causal paths, start at the exposure (osteoporosis medication), contain only arrows pointing away from the exposure, and end at the outcome (cardiovascular disease). Meanwhile, the black lines are biasing paths closed by restriction of the study participants (i.e., those with osteoporosis or experiencing fragility fractures) or adjustment. Facility care structure includes number of hospital beds and function classified according to the Diagnosis Procedure Combination medical institution group classification.

Abbreviations; CVD: cardiovascular disease; Af: atrial fibrillation; CKD: chronic kidney disease; RA: rheumatoid arthritis; DM: diabetes mellitus; COPD: chronic obstructive pulmonary disease; BMI: body mass index.

**eMethods 1. Definition of outcomes of the present study**

Myocardial infarction (MI) was detected by an algorithm that analyzes the diagnosis code (ICD-10), hospitalization according to the claim data, and an identification of accompanying treatment (i.e., either percutaneous coronary intervention, coronary artery bypass grafting, intra-aortic balloon pumping, percutaneous cardiopulmonary support, or thrombolytic therapy) as indicated by the medical procedure code or drug code (WHO-ATC code). These treatments must be provided within 30 days of the admission for MI.^1,2^

Cerebral infarction (CI) was detected by an algorithm that analyzes the diagnosis code (ICD-10), an imaging test (at least one of computed tomography, magnetic resonance imaging, or magnetic resonance angiography performed within 30 days of the diagnosis code), hospitalization according to the claim data, and an identification of accompanying treatment (i.e., craniotomy, mechanical thrombectomy, cerebroprotective agents, intravenous antiplatelet agents, intravenous anticoagulant agents, thrombolytic agents, or anti-edema agents) as indicated by the medical procedure code or drug code (WHO-ATC code). This treatment must be provided within 30 days of the admission for CI.^3,4^

Cerebral hemorrhage (CH) was detected by an algorithm that analyzes the diagnosis code (ICD-10), an imaging test (at least one of computed tomography, magnetic resonance imaging, or magnetic resonance angiography observed within 30 days of the diagnosis code), hospitalization according to the claim data, and an identification of accompanying treatment (i.e., hematoma removal, anti-edema agents, or intravenous antihypertensive agents) as indicated by the medical procedure code or drug code (WHO-ATC code). These treatments must be provided within 30 days of the admission for the CH. ^3,4^

**eMethods 2. Detailed description of why observations with a facility-level prescription proportion of zero were excluded**

In the first stage of the two-stage residual inclusion procedure, we assume the following model:^1^

$$A=\mathrm{expit} \left( \beta_{0}+\beta_{1}IV+\sum_{k=2}^{p} \beta_{k}X_{k} \right)+U$$

where *A* is the treatment, *IV* is the instrumental variable, *X_k_* (*k* = 2,…,*p*) are covariates, *U* is the unmeasured confounder assumed to be uncorrelated with *IV* and has zero expectation, and expit(*k*) = 1/(1+exp(−*k*)). Under this model, we have

$$R=A-\mathrm{expit} \left( \beta_{0}+\beta_{1}IV+\sum_{k=2}^{p} \beta_{k}X_{k} \right)=U.$$

This formula indicates that the first stage residual *R* is identical to the unmeasured confounder. However, if the probability of *A* = 1 is 0, i.e., $\Pr\left( A=1 | IV, X \right)=expit \left( \beta_{0}+\beta_{1}IV+\sum_{k=2}^{p} \beta_{k}X_{k} \right)$ = 0, *R* is also equal to zero. Thus, in this case, we cannot predict the value of $U$ by *R*. This is why observations with a facility-level prescription proportion of zero were excluded.
